# Development of minimum reporting sets of patient characteristics in epidemiological research: a methodological systematic review

**DOI:** 10.1101/2023.02.07.23285508

**Authors:** My Luong Vuong, Pham Hien Trang Tu, Khanh Linh Duong, Tat-Thang Vo

## Abstract

**Background:** Core patient characteristic sets (CPCS) are increasingly developed to identify variables that should be reported to describe the target population of epidemiological studies in the same medical area, while keeping the additional burden on the data collection acceptable.

**Methods:** We conduct a systematic review of primary studies/ protocols published aiming to develop CPCS, using the PubMed database. We particularly focus on the study design and the characteristics of the proposed CPCS. Quality of Delphi studies was assessed by a tool prosposed in the literatue. All results are reported descriptively.

**Results:** Among 23 eligible studies, Delphi survey is the most frequently used technique to obtain consensus in CPCS development (69.6%, n=16). Most studies do not include patients as stakeholders. The final CPCS rarely include socioeconomic factors. 60.9% (n=14) and 31.6% (n=6) of studies provide definition and recommend measurement methods for items, respectively.

**Conclusion:** This study identified a considerable variation and suboptimality in many methodological aspects of CPCS studies. To enhance the credibility and adoption of CPCS, a standard for conducting and reporting CPCS studies is warranted.

**Funding:** No funds, grants, or other support were received during the preparation of this manuscript.

**Registration:** This review was not pre-registered.

## Introduction

In epidemiological research, collecting and reporting patient characteristics are of crucial importance. These data allow one to assess the generalizability (or external validity) of the obtained findings, by looking at how closely the study samples match patients in a realistic healthcare setting (1). When comprehensive patient characteristics data are available, the difference between a study sample and a clinically relevant patient population can even be statistically accounted for, to improve the applicability of the findings in clinical practice (2).

Beyond external validity, patient characteristic data is also helpful to improve internal validity. For instance, by assessing the balance of important outcome prognostic factors across different treatment groups in a trial, one can assess whether there might be imperfect randomization. This is highly important when trials are of small sample size (such as in cancerology, where algorithms like minimization-based methods are often used to determine the treatment assignment for each patient) or trials with specific design (such as clusters randomized) (3, 4). In pragmatic trials, detailed patient characteristic data is also strongly needed to account for adherence and drop-out, especially when the aim is to estimate per-protocol treatment effects or to handle missing data (5). Likewise, in observational studies, assessing the balance of exposure and non-exposure groups after propensity score-based stratification or matching, for instance, require extensive data on patient characteristics (6).

In systematic reviews and evidence synthesis, when the eligible studies collect and report data on a common set of patient characteristics, the assessment of the target population (factor P in the PICO criteria) across studies will be facilitated. A more insightful evaluation of the heterogeneity observed among trial results will also be possible (7, 8). Recently, novel methods for causally interpretable meta-analysis have been proposed (8–11). These frameworks also rely on having a rich set of (prognostic) patient characteristics collected across individual studies.

Despite its importance in practice, the collection and reporting of patient characteristic data remains inconsistent and suboptimal. Cahan et al (2017) recently showed that among 186,941 trials on ClinicalTrials.gov, only 8.9% reported baseline participant measures, and up to 85% of those measures were reported only once in the entire registry (12). Lack of adequate reporting of important prognostic factors was also highlighted by Wertli et al. (2013), when they assessed 84 low back pain trials and found that almost half of them incompletely reported variables that are of prognostic importance, even with easily obtainable variables such as age or comorbidities (13). Similar issues also prevalent in many other medical fields, including asthma, diabetes, hypertension, or colorectal cancer (14–18).

In these recent years, significant efforts have been made to standardize the collection and reporting of patient characteristics in epidemiological research. Across many therapeutic areas, a so-called core patient characteristic set (CPCS) is specifically developed to identify all key prognostic factors that should be commonly collected and reported (among studies and databases evaluating a target medical condition), while keeping the additional burden on the implementation acceptable (**Fig. 1**). Above and beyond the variables proposed in the core set, researchers are free to measure and report additional patient characteristics that are of relevance to their topic. This CPCS concept is inspired by (and hence closely related to) the concept of core outcome set (COS) proposed in clinical research. However, while the methodology for COS development is increasingly enriched in the literature, little attention has been given so far to CPCS and how to develop it in practice.

**Fig. 1.**
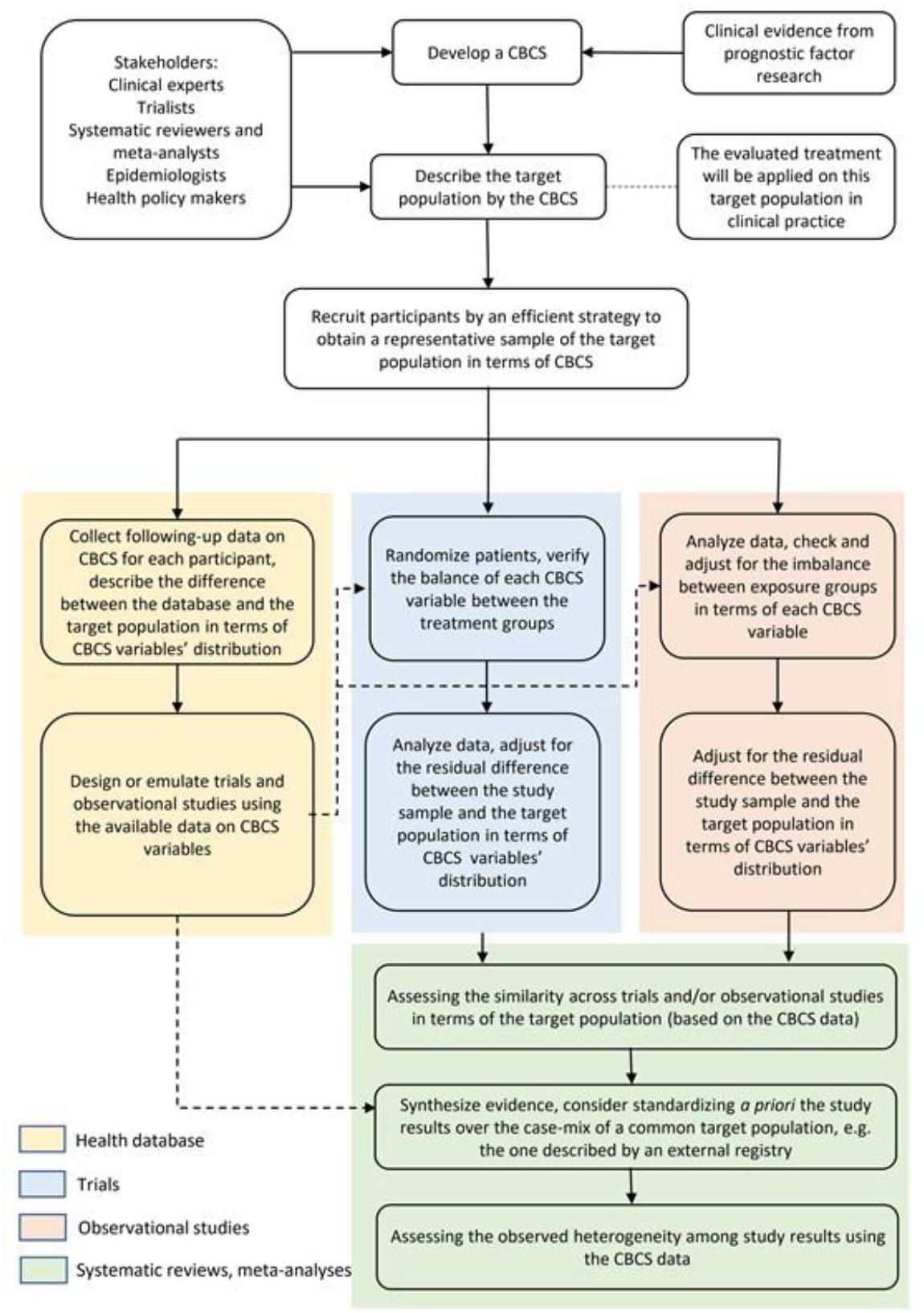
The use of core patient characteristic sets in epidemiological research

In this paper, we aim to describe the methodology of studies aiming to establish a core set of patient characteristics that should be commonly measured and reported in epidemiological studies and/or in large medical cohorts. By shedding light on current practice and challenges in CPCS development, this review could pave the way for future recommendations and guidelines on methodological standards of CPCS, thus enhancing the adoption of this concept in epidemiological research.

## Methods

### Study design

We conduct a methodological systematic review conforming to Preferred Reporting Items for Systematic Reviews and Meta-Analyses (PRISMA) 2020 statement (19).

### Eligibility criteria

We include primary studies or study protocols that aim to establish a core set of patient characteristics that should be commonly measured and reported in epidemiological studies and/or databases of a pre-specified medical condition, published between 01/01/2001 and 11/08/2022. We thus exclude studies that establish patient characteristics sets for other purposes such as to guide therapeutic decision-making in clinical practice. Conference abstracts, editorials, commentaries, and letters to the editor are excluded. Non-English publications and articles without full text accessible are also excluded from our review.

### Search strategy

A structured search in the PubMed database is undertaken by P.H.T. Tu on 12/08/2022. The full search strategy is available in Appendix S1. This search strategy is first developed by two reviewers (P.H.T. Tu and K.L. Duong), then further optimized by a senior researcher (T.-T. Vo) and a librarian specialized in epidemiological systematic reviews. We also manually screened the reference lists of the eligible articles to identify additional eligible studies.

### Study selection

The search results are downloaded into Endnote and imported into Rayyan web-based software (20). Duplicates are removed by the duplicate search function in Endnote and by manually reviewing the records list. Four reviewers (P.H.T. Tu, K.L. Duong, M.L. Vuong and T.H.T. Nguyen) independently screen titles and abstracts of retrieved records to select eligible papers based on the inclusion criteria. Each reviewer screens 25% of the total number of records and double-checks 20% of the work of another reviewer. Disagreements are resolved by discussion among four reviewers, and consultation with a senior researcher (T.-T. Vo).

### Data extraction and assessment

The data extraction form is developed by M.L. Vuong and P.H.T. Tu, pilot-tested and refined by K.L. Duong and T.-T. Vo (Appendix S2). The structure of the data extraction form is partially adapted from Boulkedid et al (2011) and Diamond et al (2014) (21, 22). Data extraction is performed by M.L. Vuong, P.H.T. Tu and K.L. Duong. Each reviewer extracts 33% and double-checks 33% of the total number of records. Any discrepancy is resolved by discussing among the three reviewers.

We extract the following information from the eligible studies: [1] publication year, [2] target medical conditions, [3] purposes of the developed CPCS (to use in epidemiological studies or in registry settings), [4] study design (consensus-reaching or non-consensus methods), and [5] geographical scope of the study (international or national-wide).

As Delphi technique is the most frequently used method among the eligible studies, we evaluate the methodological and reporting quality of Delphi studies with greater thoroughness. The following characteristics of Delphi studies are extracted: [1] study participants (number, response rate, types, selection criteria of participants, and whether authors report how representativeness of participants is ensured), [2] method to establish the primary list of items before Delphi rounds, [3.1] questionnaire round characteristics: number of rounds, purpose of each round, questions formulation (rating scale or open question), whether the rating scale (if used) is well-defined (i.e. number and the meaning of levels in the scales are specified), whether the questionnaire’s content is publicly available and is piloted in advance, summary information sent to respondents after each round, and methods used to encourage participants to complete the questionnaires, [3.2] in-person meetings characteristics: number of meetings and purposes, form of rating scale (if used) and whether the rating scale is well-defined, whether participants from questionnaire rounds are all invited to the meetings or only selectively, and the timing of meetings, [3.3] whether new items are allowed to be added between rounds, and [4] how consensus are defined and attained, and how Delphi process is terminated.

In the absence of a standardized, validated quality scores for Delphi studies, we roughly assess the quality of those studies by using the checklist proposed by Diamond et al (2014) (22). Four items in the checklists include [1] the reproducibility of criteria for participant selection and whether the number of Delphi rounds, [3] the criteria for dropping items at each round and [4] the criteria to stop the Delphi process are stated and prespecified. The number of items satisfied in each Delphi study is then reported as quality score. Three reviewers (M.L. Vuong, P.H.T. Tu and K.L. Duong) independently assess the quality of all Delphi studies by this tool and reach final consensus.

For the remaining studies, we narratively describe the study design, number and type of participants and organization among them, and method to establish the final CPCS. With non-Delphi, consensus-reaching study, we also extract information on method to establish the primary list of items and the definition and attainment of consensus.

Finally, we extract details of the final CPCS obtained. These include, [1] whether description of item flow reported, [2] whether only the final set or also intermediate results were reported, [3] whether the items in the final set were ranked and how, [4] number of items in the final set, [5] whether the definition and measurement of included items were given and [6] domains of items in the CPCS (demographic, clinical, patient history, socioeconomic or healthcare setting factors).

### Data synthesis

Continuous variables were presented with median and interquartile range. Categorical variables were summarized with frequencies and percentages. To investigate the content pattern of the final lists of items across eligible studies, we performed a hierarchical, complete-linkage clustering analysis (23). For each final list, we first calculate the percentage of each domain. The domain profile for each study was then used to calculate the matrix of between-study Euclidean distances. Finally, the obtained result was visualized by a tree-structure graphic.

Data analysis was performed using Microsoft Excel 365 and R version 4.1.1.

## Results

### Study selection

The PRISMA flow diagram summarizing the screening process is presented in **Fig. 2**. Of all 5819 references identified, 23 articles met the inclusion criteria for review.

**Fig. 2.**
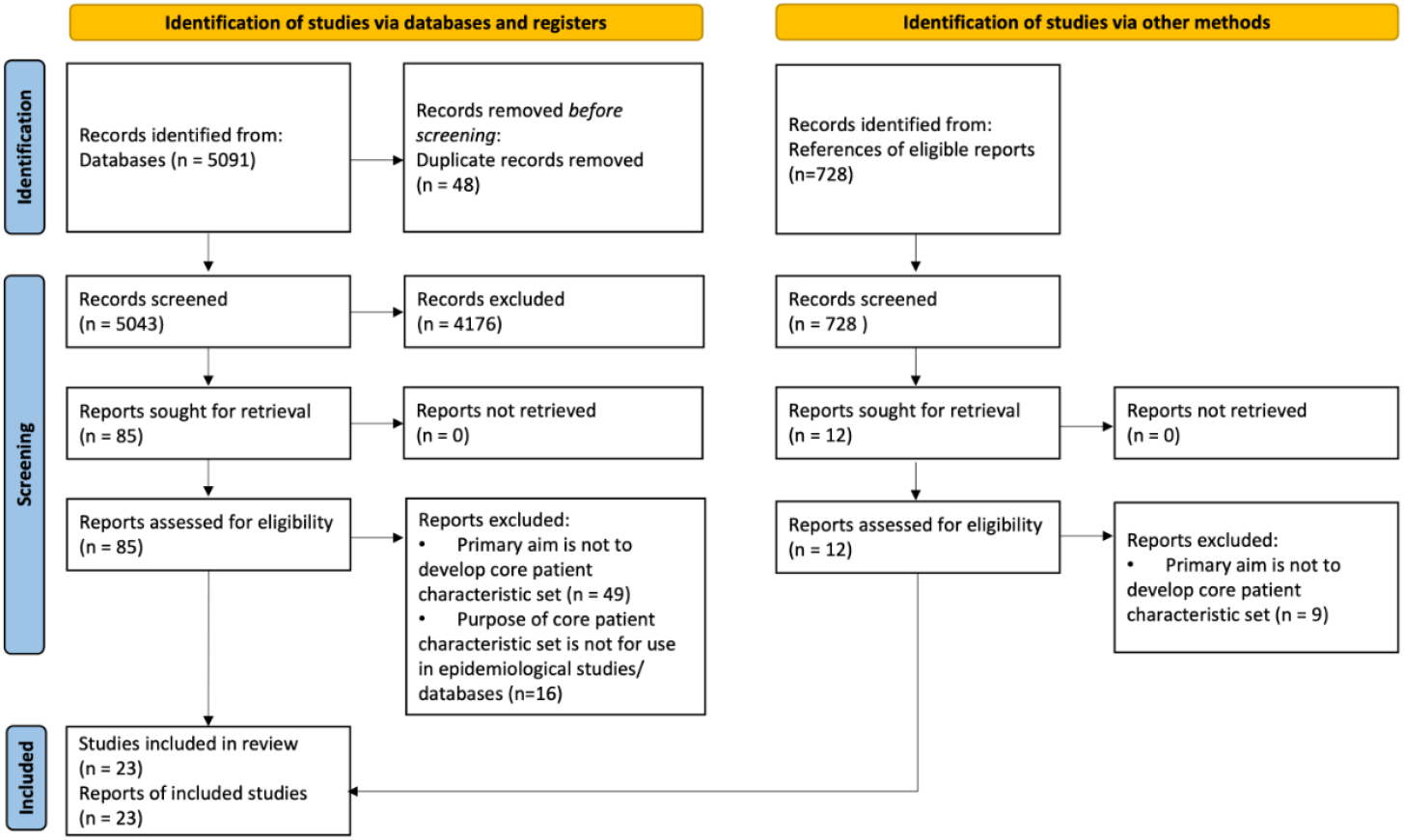
Study selection PRISMA Flowchart. PRISMA: Preferred Reporting Items for Systematic Reviews and Meta-Analyses

### General characteristic of included studies

The general characteristics of all the studies included (24–42) are provided in Table 1. Among 23 eligible studies, 73.9% (n=17) develop a CPCS for epidemiological studies, 21.7% (n=5) develop such a set for heathcare registries and one study (4.3%) develops a CPCS for both registries and epidemiological studies. About 91% of studies (n=21) were published in the last ten years, and 78.3% of studies (n=18) has an international scope. Regarding methodology, 87.0% of studies (n=20) consider a consensus reaching method to develop the core set, with Delphi being the most frequently used technique (69.6%, n=16). Other non-consensus methods include systematic review (8.7%, n=2) and conceptual analysis (4.3%, n=1).

**Table 1.** General characteristics of eligible studies (N=23)

| Study characteristics | N | % |
| --- | --- | --- |
| Publication year |  |  |
| - Before 2010 | 2 | 8.7 |
| - 2011-2015 | 5 | 21.7 |
| - 2016-2021 | 16 | 69.6 |
| Target medical conditions |  |  |
| - Circulatory system | 5 | 21.7 |
| - Oncology | 4 | 17.4 |
| - Pediatrics | 3 | 13.0 |
| - Musculoskeletal system and connective tissue | 2 | 8.7 |
| - General symptoms and signs | 2 | 8.7 |
| - Medical intervention: laparoscopic hysterectomy | 2 | 8.7 |
| - Others <sup>a</sup> | 4 | 21.7 |
| Purposes: Improving patient characteristics reporting in: |  |  |
| Epidemiological studies: | 17 | 73.9 |
| - Clinical trials | 8 | 34.8 |
| - Both clinical trials and observational studies | 9 | 39.1 |
| Registries or healthcare programs | 5 | 21.7 |
| Both registry and epidemiological studies | 1 | 4.3 |
| Study designs |  |  |
| Consensus-reaching methods | 20 | 87.0 |
| - Traditional Delphi | 8 | 34.8 |
| - Modified Delphi | 8 | 34.8 |
| - Non-Delphi | 4 | 17.4 |
| Non-consensus-reaching methods | 3 | 13.0 |
| - Systematic review | 2 | 8.7 |
| - Conceptual analysis | 1 | 4.3 |
| Geographic scopes of study participants |  |  |
| - International | 18 | 78.3 |
| - National-wide | 1 | 4.3 |
| - Not reported | 4 | 17.4 |

<sup>a</sup> Including burns, chronic fatigue syndrome, rehabilitation, and hemophilia.

### Methodological characteristics of Delphi studies

The methodological characteristics of 16 eligible Delphi studies are provided in Table 2 and Appendix S3. Remarkably, almost all studies involve healthcare professionals (93.8%, n=15) or researchers (81.3%, n=13), while only one study (8.3%) involves patients or patient representatives. The criteria for selecting participants are quite various across studies, but most commonly based on scientific renown, publishing and/or expertise level (58.3%, n=7). Though the acceptance rate of the eligible studies is relatively low (median of 25 participants versus 40 invitations), only 41.7% of studies (n=5) reported how they ensured the representativeness of participants.

**Table 2.** Methodological characteristics of studies using Delphi consensus approaches (N=16)

| **Methodological characteristics** | **N** | **%** |
| --- | --- | --- |
| **1. Participants** |  |  |
| Type of stakeholders<sup>a</sup> |  |  |
| • Healthcare professionals | 15 | 93.8 |
| • Researchers, including clinical trialists, epidemiologists, statisticians, and/or public health experts | 13 | 81.3 |
| • Data or registry/program managers | 3 | 18.8 |
| • Policy makers | 2 | 12.5 |
| • Insurance experts | 2 | 12.5 |
| • Patient representatives | 1 | 6.3 |
| The study reports how representativeness of participants is ensured | 6 | 37.5 |
| **2. Delphi rounds** |  |  |
| **2.1. Questionnaire rounds (in all 16 Delphi studies)** |  |  |
| Number of rounds, median [IQR] | 2.5 | [2–4] |
| Question formulation |  |  |
| • Rating scale for each item | 16 | 100.0 |
| ○ Binary scale | 2 | 12.5 |
| ○ Likert scale | 3 | 18.8 |
| ○ Unclear scale format | 1 | 6.3 |
| • Open questions in addition to rating scale | 10 | 62.5 |
| Rating scale/score well defined | 14 | 87.5 |
| **2.2. In-person/teleconference meeting rounds (in 8 modified Delphi studies)** |  |  |
| Number of rounds, median [IQR] | 2 | [1–3] |
| Rating scale formulation | 7 | 43.8 |
| • Binary scale | 5 | 31.3 |
| • Six-point Likert scale | 1 | 6.3 |
| • Unclear scale format | 1 | 6.3 |
| Rating scale/ score well defined | 4 | 25.0 |
| **3. Consensus definition and attainment** |  |  |
| Criteria for selecting/dropping items at each round based on: |  |  |
| • (i) Pre-defined cut-off(s) of % of participants voting certain rating level(s) | 9 | 56.3 |
| • (ii) Pre-defined cut-off(s) of a median score on a rating scale | 1 | 6.3 |
| • Both (i) and (ii) | 4 | 25.0 |
| • Unclear/ not reported | 2 | 12.5 |
| Reason to terminate Delphi process |  |  |
| • After completing the number of rounds prespecified | 14 | 87.5 |
| • When consensus is reached | 1 | 6.3 |
| • Unclear | 1 | 6.3 |

<sup>a</sup> Each study may be classified in more than one category; IQR: interquartile range.

Across all studies (100%, n=12), rating scales are used to judge the importance of items during the questionnaire rounds. These scales range from two-point to ten-point, with five-point scales being the most commonly used (33.3%, n=4). The scale is deemed as well-defined in 83.3% of studies (n=10). Apart from item rating, open-ended questions are also included in 66.7% of studies (n=8), mostly to collect qualitative feedback from participants (58.3%, n=7). Besides, 41.7% of studies (n=5) report the use of a specific method to encourage participants to complete the questionnaires (e.g., by sending them reminders or vouchers).

In six (modified) Delphi studies, in-person meetings or teleconferences are additionally organized. The median number of meetings is two (IQR 1–4). The aim of these meetings is to have discussions among participants before rerating the existing items (41.6%, n=5) and adding new items (8.3%, n=1). The rating scales used in these meeting rounds are mainly binary scales (25.0%, n=3), and are well-defined in four studies (33.3%). Meetings are scheduled at different timepoints, either before (8.3%, n=1), in between (25.0%, n=3) or after the questionnaire rounds (16.7%, n=2).

Finally, 16.7% of studies (n=2) do not report the criteria for selecting or dropping an item (Appendix S3). In 91.7% of studies (n=11), the Delphi process is terminated when the preplanned rounds are completed, regardless of the stability of responses or whether consensus has been obtained for all items. In one study, the reason for termination is unclear. As stopping the Delphi not based on response stability or consensus is deemed as suboptimal (22), all studies are penalized for this in the subsequent quality assessment. More precisely, 50% (n=6) of studies have a quality score of three, and 50% (n=6) of studies have a quality score of one or two, on the four-point quality scoring system proposed by Diamond et al. (2014) (22) (Appendix S3).

### Methodological characteristics of non-Delphi studies

The methodological characteristics of seven non-Delphi studies are provided in Table 3. In general, only one study (14.3%) reports the types of stakeholders participating in the construction of the CPCS, and no studies report number nor distribution of stakeholders. Similarly, no studies report the criteria for selecting/dropping each item, neither how consensus is reached after each round and at the end.

**Table 3.** Other studies of non-Delphi methods for core patient characteristic set (CPCS) construction

| No | Study | Study design | Settings | Participants | Methodology |
| --- | --- | --- | --- | --- | --- |
| Consensus-reaching methods |  |  |  |  |  |
| 1 | Jones 2018 | NDC | Core data element to promote interoperability across registries, clinical care and trials of peripheral vascular intervention (PVI) | Type of stakeholders<br>- Healthcare professionals<br>- Researchers<br>- Policy makers<br>- Health information technology vendor<br>- Study device manufacturers.<br>Number and distribution of stakeholders<br>- Not reported.<br>Organization among stakeholders:<br>- Clinical group: identify core clinical concepts.<br>- Informatics group: develop clinical concepts into data elements.<br>- Device identifier database group: facilitate the use of device data. | Method to establish primary list of items<br>- Informatics group extracts data elements from existing registry data forms and device company case report forms and selects data elements that are deemed specific to peripheral vascular disease.<br>Method to reach consensus<br>- Interactive web conferences and face-to-face meetings are organized among clinical group members to reach consensus on key data elements.<br>- The included elements were prioritized based on their availability in existing data sources and applicability in PVI clinical studies.<br>Criteria for selecting/ dropping items and consensus attainment:<br>- Not reported. |
| 2 | Storrow 2012 | NDC | Guideline for reporting criteria in evaluation and management studies on acute heart failure syndromes | Type of stakeholders<br>- Not reported.<br>Number and distribution of stakeholders:<br>- Not reported.<br>Organization among stakeholders<br>- A working group is in charge of developing and finalizing the guideline.<br>- External experts from two specialized organizations are invited to review the guideline. | Method to establish primary list of items<br>- Based on existing clinical guidelines<br>Method to reach consensus<br>- At in-person meetings, each element in the list was discussed among the working group members for incorporation as a core measure, supplemental measure, or dropped from further consideration.<br>- The draft guideline was then distributed to external experts in two specialized organizations to review, revise, and finalize before publication. |
|  |  |  |  |  | Criteria for selecting/ dropping items and consensus attainment:<br>- Not reported. |
| 3 | Kwakkel 2017 | NDC | Core measurement set in stroke rehabilitation and recovery trials | Type of stakeholders<br>- International multidisciplinary stroke experts.<br>Number and distribution of stakeholders:<br>- Around 60 experts.<br>Organization among stakeholders<br>- All stakeholders are included in a working group. | Method to establish primary list of items.<br>- Based on a literature review by a small group of three to five members.<br>Method to reach consensus on the final list.<br>- Data are presented to the working group at the meeting and discussed after meeting, no formal consensus approach employed due to shortness of time.<br>Criteria for selecting/ dropping items and consensus attainment:<br>- Not reported. |
| 4 | Jason 2012 | NDC | Minimum data elements in chronic fatigue syndrome research reports | Type of stakeholders<br>- Unclear.<br>Number and distribution of stakeholders:<br>- Not reported.<br>Organization among stakeholders<br>- Not applicable. | Not reported. |
| Non consensus methods |  |  |  |  |  |
| 5 | Driessen 2016 | SR | Minimal set of potential case-mix variables for studies in laparoscopic hysterectomy | Not applicable | Method to establish the final list:<br>- Patient characteristics that significantly predict the study outcome are extracted from eligible studies.<br>- Case-mix characteristics were stratified by level of evidence (based on methodological quality score of corresponding studies, assessed using Newcastle-Ottawa Quality Assessment Scale). |
| 6 | Osooli 2015 | SR | Minimal data set for registries of hemophilia | Not applicable | Method to establish the final list:<br>- Predictors of study outcome were extracted from registry-based studies.<br>- Time of reporting and statistical significance of each variable among the registries are reported.<br>- Criteria to finalize the list are not reported. |
| 7 | Meyer 2020 | CA | Reporting standards of patients' characteristics in rehabilitation trials | Type of stakeholders<br>- Authors and participant of a rehabilitation methodology meeting.<br>Number and distribution of stakeholders:<br>- Not reported.<br>Organization among stakeholders<br>- Main author: conduct analysis and prepare drafts.<br>- Co-authors and meeting participants: revise the drafts. | - The main author identified core patient-specific characteristics based on the concept of target condition definition, subjects' characteristics, and service description.<br>- The first and third drafts of the analysis were revised by the participants, and the second draft was revised by co-authors. |

NDC: non-Delphi consensus, SR: systematic review, CA: conceptual analysis

### Characteristics of the final lists of patient characteristics

The reporting of results and characteristics of the final CPCSs are provided in Table 4 and **Fig. 3**. Almost all studies (91.3%, n=21) report the final CPCS. Studies that develop a CPCS for in registries often have more items than those developing a CPCS for epidemiological studies (26 [10-31] vs 17 [10-23]) (Table 4).

**Table 4.** The reporting of results among all eligible studies (N=23)

| Characteristics | Target of the CPCS, N (%) |  | All studies<br>N (%)<br>(N=23) |
| --- | --- | --- | --- |
|  | Epidemiological<br>studies (N=17) | Registries<br>(N=6) |  |
| CPCS details reported | 16 (94.1) | 5 (83.3) | 21 (91.3) |
| Number of items in the CPCS, median [IQR] | 17 [10–23] | 26 [10–31] | 17 [10–25] |
| Priority of items in the CPCS |  |  |  |
| • Based on level of consensus | 3 (17.6) | 1 (16.7) | 4 (17.4) |
| • Based on level of detail/ complexity | 3 (17.6) | 1 (16.7) | 4 (17.4) |
| • Based on level of evidence | 1 (5.9) | - | 1 (4.3) |
| • Unclear | 2 (11.8) | - | 2 (8.7) |
| • Not considered | 8 (47.1) | 4 (66.7) | 12 (52.2) |
| Items in the CPCS defined | 10 (58.8) | 4 (66.7) | 14 (60.9) |
| Measurement of non-obvious items in the CPCS |  |  |  |
| • Units | 4 (23.5) | 4 (66.7) | 8 (34.8) |
| • Measurement methods | 2 (11.8) | 4 (66.7) | 6 (26.1) |

IQR: interquartile range

**Fig. 3.**
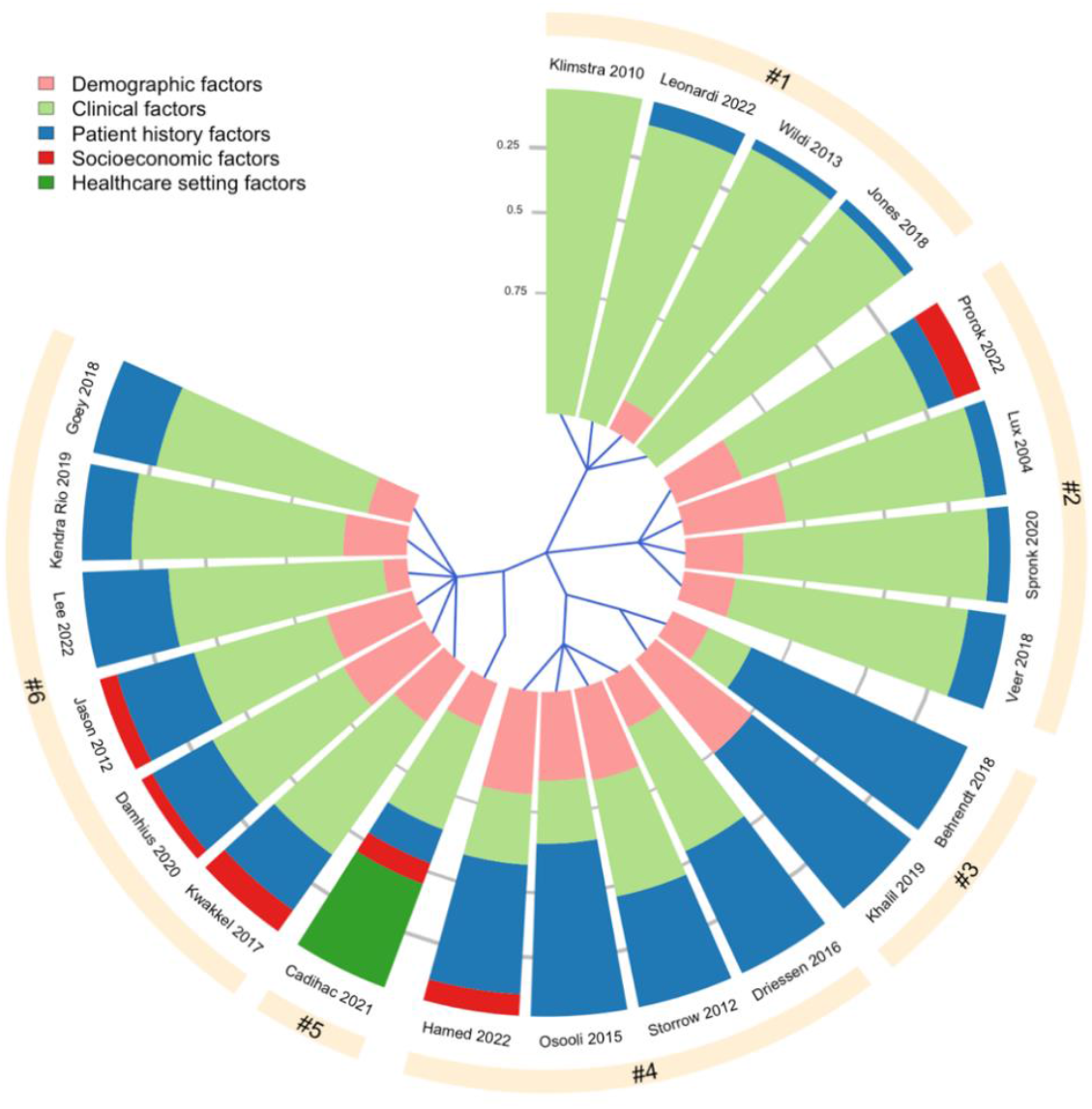
Hierarchical clustering of 21/23 CPCS based on five variable domains, namely [1] Demographic factors (age, gender, race), [2] Clinical factors (e.g., disease severity, signs and symptoms, laboratory test), [3] Patient history factors (e.g., lifestyle factors, comorbidities, family history), [4] Socioeconomic factors (e.g., level of education, income, occupation), [5] Healthcare setting factors (e.g., ambulatory care, inpatient, or ICU). Each slice of the chart represents one CPCS.

The characteristics of the final CPCSs is provided in **Fig. 3**. Most CPCSs contain demographic factors (age, gender, race), clinical factors (e.g., disease severity, presence of a symptom, laboratory test) and patient history factors (e.g., lifestyle, comorbidities, family history). In contrast, socioeconomic factors and healthcare settings factors are often absent in most final lists.

Items included are defined in 60.9% of CPCS (n=14). Besides, 34.8% (n=8) and 26.1% (n=6) of CPCS have specific recommendations on the scale and measurement method for non-obvious items, respectively (Table 4).

The sectors in each chart indicate what type of variables were included in each CPCS, with the area of each sector corresponds to the proportion of each variable type within one CPCS. For instance, the CPCS developed by Khalil et al. (2019) consists of two variable domains: demographic factors and patient history factors, which make up 25% and 75% of the CPCS, respectively. The blue lines starting from the center of the chart define how the tools are divided into the six clusters. Clusters #3 and #4, and #5 and #6 are grouped as sub-nodes of two major nodes, meaning that the tools in these sub-nodes have more similar domain profile compared to the tools in other clusters.

## Discussion

The call for better patient characteristics collection and reporting in epidemiological research is not new. The Consolidated Standards of Reporting Trials (CONSORT) statement is one of the first initiatives aiming to improve the reporting of trials, including the selection criteria (item 4a) and the description of the resulting samples (item 15). A table showing baseline demographic and clinical characteristics for each treatment group, including the baseline measurement of the outcome is required. However, the CONSORT statement provides no further indication of which patient characteristics to report. Extensions of the CONSORT statement specify that information on socioeconomic variables should be added, and that all relevant prognostic variables should be reported, but only one CONSORT extension explicitly asks to include comorbidity. Another initiative is the Food and Drug Administration Amendments Act (FDAAA) mandates which require all covered studies to report results (including participants’ age, gender, race or ethnicity, and the baseline measures of the primary outcome) within 1 year of completion (43).

Constructing core patient characteristics sets is increasingly considered as a new method to further improve the collection and reporting of patient characteristics. Most CPCSs are developed within the last ten years. Not only for improving internal and external validity of epidemiological studies, many CPCSs are also developed to increase the quality of patient characteristics data in registries. This is essential because registries are becoming important data sources for recent epidemiological research.

In this review, we identify many different methods to construct a CPCS. Among these methods, consensus-reaching techniques such as Delphi survey are the most frequently used. Indeed, Delphi survey is one of the ideal methods to collect expert-based judgements when the available knowledge is incomplete, which is often the case in CPCS or COS development (44).

Most Delphi studies in our review do not include patients as stakeholders. This is probably because CPCS development requires specialized knowledge on prognostic factors of a certain disease. Hence, involving patients would bring little benefit to the process. However, embracing patients’ perspective on certain variables in the final set could be helpful, especially when these variables are private information of patients such as socioeconomic status, income, family history etc. Methods for patient engagement has recently been proposed for core outcome set, which could be further adjusted for the development of CPCS (45, 46). Besides, many CPCS studies do not report how the representativeness of participants is ensured. Such information is important to determine the quality of the obtained CPCS and its uptake, hence should be better reported in future practice.

Our review has identified a wide range of consensus definitions employed by Delphi studies, with the most common definitions based on the pre-defined cut-offs of percentage of participants voting certain rating levels. This is in line with findings from previous reviews (22, 47). Earlier studies also acknowledged the difficulty of ascertaining the validity of consensus definitions, and there has been no specific guidance on best consensus definition method, which could explain the observed variability in our study (22). However, the minimum standard is to report comprehensibly how consensus is defined and achieved throughout the process. This is not satisfied by one-sixth of eligible studies. Lack of clarity on this could render the studies susceptible to bias and arbitrariness during data collection, analysis and interpretation (47).

Most of studies stop the Delphi process after completing a prespecified number of rounds, regardless of the consensus attainment status. Considering the scarcity and/or divergence of evidence, perfect consensus for 100% of items may not be achievable. Indeed, it has been shown that the evidence of many prognostic variables greatly suffers from a high risk of publication bias, selective reporting biases, poor statistical analyses and so forth (48). To compromise on this issue, many CPCS studies choose to group items into different sets with different priority (based on level of evidence and/or consensus), so that researchers will also be informed about the quality of the variables in the final set. On the other hand, it is important to update the CPCS over time when novel evidence for new (and current) prognostic factors are available in the literature.

Regarding non-Delphi studies, the methodological reporting is relatively weak. Many important factors such as characteristics of experts, method to establish the final list and consensus attainment were often not reported. This raises concern about the rigor of the CPCSs obtained from these studies.

Our review also provides many important remarks on the final core sets across studies. First, while demographic, clinical and patient history factors are dominant in all final sets, socioeconomic and healthcare setting factors are often overlooked. This is suboptimal. Indeed, the socioeconomic gradient in health is ubiquitous, and has been described across pathologies, in life expectancy and mortality (49–51). Meanwhile, describing the healthcare setting is important to assess the applicability of any epidemiological findings into practice. These two types of factors are not any less important than other clinical factors often included in the CPCSs.

Second, the number of (final) items in CPCSs for registries is often higher than in CPCS for epidemiological studies. This could be because registries are of large scale and have more (financial and human) resources for data collection than in traditional epidemiological research (41). The disparity between CPCS for registries and epidemiological studies, however, could imply the challenges in the interoperability between these two settings, and the adoption of CPCS from one setting to the other within one medical field.

Finally, apart from a list of important patient characteristics to collect and report, many CPCSs also provide recommendations on the measurement methods and scales for non-obvious or subjective items. Doing so could further reduce the heterogeneity and inconsistency in data collection practice, as the conversion between different scales for many variables is not straightforward. The downside of this, however, could be that the applicability of the proposed CPCS is reduced in practice. For instance, the recommended measurement methods might be not widely used or have a high cost. These practical factors should be taken into account when making recommendations on the core set.

It is important to acknowledge some limitations of our study. First, we limited the eligibility criteria to articles published in English, hence appropriate studies that were not published in English might have been excluded. Second, the great difference between the number of records identified from the literature and the number of eligible studies may arise from the fact that the specificity and coverage of our search strategy is not optimal. Such a challenge stems from the discrepancies and non-standardized terminology for CPCS, as opposed to COS. We mitigate this issue by consulting a librarian specialized in epidemiological systematic reviews to optimize the search strategy, and by manually search for additional eligible studies from the reference list of identified eligible studies. Finally, we were not able to conduct a formal quality assessment for Delphi studies nor CPCS studies in general, as tools for such purpose are not yet available in the literature.

## Conclusion

This methodological systematic review has revealed the suboptimality of the conduct and reporting of CPCS studies, particularly in participant characteristics, method to obtain the final CPCS, and coverage and detail of the CPCS obtained. A conduct and reporting standard for CPCS studies is thus warranted, to further enhance the quality of CPCSs and promote the adoption of this concept in epidemiological research.

## Data Availability

All data produced in the present study are available upon reasonable request to the authors

https://1drv.ms/u/s!AnSwAV_ksnYHgvF5TkCdP4565VCm0w?e=EPt0gP

## Declarations

### Ethics approval

Our study results were based on previously published data, none of which is stated as research involving human or animal subjects. Ethical approval is thus not required.

### Data and computing code availability

All data extracted for the systematic review and R code are made available as supplementary information (Appendix S4&S5).

### Funding

*The authors received no financial support for the research, authorship, and/or publication of this article*.

### Registration and protocol

This review includes no clinical studies and therefore no protocol was pre-registered.

### Conflicting Interests

The Authors declare that there is no conflict of interest.

## Notes

### Competing Interest Statement

The authors have declared no competing interest.

### Funding Statement

This study did not receive any funding

## References

1. Dekkers OM, Elm Ev, Algra A, Romijn JA, Vandenbroucke JP. How to assess the external validity of therapeutic trials: a conceptual approach. International Journal of Epidemiology. 2009;39(1):89–94.

2. Lesko CR, Ackerman B, Webster-Clark M, Edwards JK. Target Validity: Bringing Treatment of External Validity in Line with Internal Validity. Current Epidemiology Reports. 2020;7(3):117–24.

3. Dickinson LM, Hosokawa P, Waxmonsky JA, Kwan BM. The problem of imbalance in cluster randomized trials and the benefits of covariate constrained randomization. Family Practice. 2021;38(3):368–71.

4. Altman DG, Bland JM. Treatment allocation by minimisation. BMJ. 2005;330(7495):843-.

5. Murray E, Swanson S, Young J, Hernán M. Guidelines for estimating causal effects in pragmatic randomized trials2019.

6. Hartz A, Marsh JL. Methodologic Issues in Observational Studies. 2003;413:33–42.

7. Debray TP, Moons KG, van Valkenhoef G, Efthimiou O, Hummel N, Groenwold RH, et al. Get real in individual participant data (IPD) meta-analysis: a review of the methodology. Res Synth Methods. 2015;6(4):293–309.

8. Vo T-T, Porcher R, Chaimani A, Vansteelandt S. A novel approach for identifying and addressing case-mix heterogeneity in individual participant data meta-analysis. Research Synthesis Methods. 2019;10(4):582–96.

9. Dahabreh IJ, Petito LC, Robertson SE, Hernán MA, Steingrimsson JA. Toward Causally Interpretable Meta-analysis: Transporting Inferences from Multiple Randomized Trials to a New Target Population. Epidemiology. 2020;31(3):334–44.

10. Manski CF. Toward Credible Patient-centered Meta-analysis. Epidemiology. 2020;31(3):345–52.

11. Sobel M, Madigan D, Wang W. Causal Inference for Meta-Analysis and Multi-Level Data Structures, with Application to Randomized Studies of Vioxx. Psychometrika. 2017;82(2):459–74.

12. Cahan A, Anand V. Second thoughts on the final rule: An analysis of baseline participant characteristics reports on ClinicalTrials.gov. PLOS ONE. 2017;12:e0185886.

13. Wertli MM, Schöb M, Brunner F, Steurer J. Incomplete reporting of baseline characteristics in clinical trials: an analysis of randomized controlled trials and systematic reviews involving patients with chronic low back pain. PLoS One. 2013;8(3):e58512.

14. Hemmingsen B, Lund SS, Gluud C, Vaag A, Almdal T, Hemmingsen C, et al. Intensive glycaemic control for patients with type 2 diabetes: systematic review with meta-analysis and trial sequential analysis of randomised clinical trials. Bmj. 2011;343:d6898.

15. Sorbye H, Köhne CH, Sargent DJ, Glimelius B. Patient characteristics and stratification in medical treatment studies for metastatic colorectal cancer: a proposal for standardization of patient characteristic reporting and stratification. Ann Oncol. 2007;18(10):1666–72.

16. van Boven FE, de Jong NW, Braunstahl G-J, Gerth van Wijk R, Arends LR. A meta-analysis of baseline characteristics in trials on mite allergen avoidance in asthmatics: room for improvement. Clinical and Translational Allergy. 2020;10(1):2.

17. van de Laar FA, Akkermans RP, van Binsbergen JJ. Limited evidence for effects of diet for type 2 diabetes from systematic reviews. European Journal of Clinical Nutrition. 2007;61(8):929–37.

18. Zhang X, Wu Y, Kang D, Wang J, Hong Q, Le P. The external validity of randomized controlled trials of hypertension within China: from the perspective of sample representation. PLoS One. 2013;8(12):e82324.

19. Page MJ, McKenzie JE, Bossuyt PM, Boutron I, Hoffmann TC, Mulrow CD, et al. The PRISMA 2020 statement: an updated guideline for reporting systematic reviews. Bmj. 2021;372:n71.

20. Ouzzani M, Hammady H, Fedorowicz Z, Elmagarmid A. Rayyan-a web and mobile app for systematic reviews. Syst Rev. 2016;5(1):210.

21. Boulkedid R, Abdoul H, Loustau M, Sibony O, Alberti C. Using and reporting the Delphi method for selecting healthcare quality indicators: a systematic review. PLoS One. 2011;6(6):e20476.

22. Diamond IR, Grant RC, Feldman BM, Pencharz PB, Ling SC, Moore AM, et al. Defining consensus: a systematic review recommends methodologic criteria for reporting of Delphi studies. J Clin Epidemiol. 2014;67(4):401–9.

23. Superchi C, González JA, Solà I, Cobo E, Hren D, Boutron I. Tools used to assess the quality of peer review reports: a methodological systematic review. BMC Medical Research Methodology. 2019;19(1):48.

24. Ahmadi M, Alipour J, Mohammadi A, Khorami F. Development a minimum data set of the information management system for burns. Burns. 2015;41(5):1092–9.

25. Damhuis SE, Bloomfield FH, Khalil A, Daly M, Ganzevoort W, Gordijn SJ. A Core Outcome Set and minimum reporting set for intervention studies in growth restriction in the NEwbOrN: the COSNEON study. Pediatr Res. 2021;89(6):1380–5.

26. Driessen SR, Sandberg EM, la Chapelle CF, Twijnstra AR, Rhemrev JP, Jansen FW. Case-Mix Variables and Predictors for Outcomes of Laparoscopic Hysterectomy: A Systematic Review. J Minim Invasive Gynecol. 2016;23(3):317–30.

27. Goey KKH, Sørbye H, Glimelius B, Adams RA, André T, Arnold D, et al. Consensus statement on essential patient characteristics in systemic treatment trials for metastatic colorectal cancer: Supported by the ARCAD Group. Eur J Cancer. 2018;100:35–45.

28. Jason LA, Unger ER, Dimitrakoff JD, Fagin AP, Houghton M, Cook DB, et al. Minimum data elements for research reports on CFS. Brain Behav Immun. 2012;26(3):401–6.

29. Jones WS, Krucoff MW, Morales P, Wilgus RW, Heath AH, Williams MF, et al. Registry Assessment of Peripheral Interventional Devices (RAPID): Registry assessment of peripheral interventional devices core data elements. J Vasc Surg. 2018;67(2):637–44.e30.

30. Khalil A, Gordijn SJ, Beune IM, Wynia K, Ganzevoort W, Figueras F, et al. Essential variables for reporting research studies on fetal growth restriction: a Delphi consensus. Ultrasound in Obstetrics & Gynecology. 2019;53(5):609–14.

31. Klimstra DS, Modlin IR, Adsay NV, Chetty R, Deshpande V, Gönen M, et al. Pathology reporting of neuroendocrine tumors: application of the Delphic consensus process to the development of a minimum pathology data set. Am J Surg Pathol. 2010;34(3):300–13.

32. Kwakkel G, Lannin NA, Borschmann K, English C, Ali M, Churilov L, et al. Standardized measurement of sensorimotor recovery in stroke trials: Consensus-based core recommendations from the Stroke Recovery and Rehabilitation Roundtable. Int J Stroke. 2017;12(5):451–61.

33. Lux AL, Osborne JP. A Proposal for Case Definitions and Outcome Measures in Studies of Infantile Spasms and West Syndrome: Consensus Statement of the West Delphi Group. Epilepsia. 2004;45(11):1416–28.

34. Meyer T, Selb M, Kiekens C, Grubišić F, Arienti C, Stucki G, et al. Toward Better Reporting Standards of Patients’ Characteristics in Rehabilitation Trials: Applying a New Conceptual Framework to Current Standards. Am J Phys Med Rehabil. 2020;99(3):216–23.

35. Osooli M, Berntorp E. Inhibitors in haemophilia: what have we learned from registries? A systematic review. J Intern Med. 2015;277(1):1–15.

36. Rio EK, Mc Auliffe S, Kuipers I, Girdwood M, Alfredson H, Bahr R, et al. ICON PART-T 2019-International Scientific Tendinopathy Symposium Consensus: recommended standards for reporting participant characteristics in tendinopathy research (PART-T). Br J Sports Med. 2020;54(11):627–30.

37. Spronk PER, Begum H, Vishwanath S, Crosbie A, Earnest A, Elder E, et al. Toward International Harmonization of Breast Implant Registries: International Collaboration of Breast Registry Activities Global Common Data Set. Plast Reconstr Surg. 2020;146(2):255–67.

38. Storrow AB, Lindsell CJ, Collins SP, Diercks DB, Filippatos GS, Hiestand BC, et al. Standardized reporting criteria for studies evaluating suspected acute heart failure syndromes in the emergency department. J Am Coll Cardiol. 2012;60(9):822–32.

39. Ter Veer E, van Rijssen LB, Besselink MG, Mali RMA, Berlin JD, Boeck S, et al. Consensus statement on mandatory measurements in pancreatic cancer trials (COMM-PACT) for systemic treatment of unresectable disease. Lancet Oncol. 2018;19(3):e151–e60.

40. Wildi LM, Hensel A, Wertli M, Michel BA, Steurer J. Relevant baseline characteristics for describing patients with knee osteoarthritis: results from a Delphi survey. BMC Musculoskelet Disord. 2013;14:369.

41. Behrendt CA, Bertges D, Eldrup N, Beck AW, Mani K, Venermo M, et al. International Consortium of Vascular Registries Consensus Recommendations for Peripheral Revascularisation Registry Data Collection. Eur J Vasc Endovasc Surg. 2018;56(2):217–37.

42. Cadilhac DA, Bagot KL, Demaerschalk BM, Hubert G, Schwamm L, Watkins CL, et al. Establishment of an internationally agreed minimum data set for acute telestroke. J Telemed Telecare. 2021;27(9):582–9.

43. Liberopoulos G, Trikalinos NA, Ioannidis JP. The elderly were under-represented in osteoarthritis clinical trials. J Clin Epidemiol. 2009;62(11):1218–23.

44. Niederberger M, Spranger J. Delphi Technique in Health Sciences: A Map. Frontiers in Public Health. 2020;8.

45. Beyer K, MacLennan SJ, Moris L, Lardas M, Mastris K, Hooker G, et al. The Key Role of Patient Involvement in the Development of Core Outcome Sets in Prostate Cancer. Eur Urol Focus. 2021;7(5):943–6.

46. Vanderhout SM, Smith M, Pallone N, Tingley K, Pugliese M, Chakraborty P, et al. Patient and family engagement in the development of core outcome sets for two rare chronic diseases in children. Research Involvement and Engagement. 2021;7(1):66.

47. Jünger S, Payne SA, Brine J, Radbruch L, Brearley SG. Guidance on Conducting and REporting DElphi Studies (CREDES) in palliative care: Recommendations based on a methodological systematic review. Palliat Med. 2017;31(8):684–706.

48. Riley RD, Hayden JA, Steyerberg EW, Moons KGM, Abrams K, Kyzas PA, et al. Prognosis Research Strategy (PROGRESS) 2: Prognostic Factor Research. PLOS Medicine. 2013;10(2):e1001380.

49. Braveman PA, Cubbin C, Egerter S, Chideya S, Marchi KS, Metzler M, et al. Socioeconomic status in health research: one size does not fit all. Jama. 2005;294(22):2879–88.

50. Kaplan GA, Keil JE. Socioeconomic factors and cardiovascular disease: a review of the literature. Circulation. 1993;88(4 Pt 1):1973–98.

51. Khalatbari-Soltani S, Cumming RC, Delpierre C, Kelly-Irving M. Importance of collecting data on socioeconomic determinants from the early stage of the COVID-19 outbreak onwards. Journal of Epidemiology and Community Health. 2020;74(8):620–3.

