## Appendix S1 for "Development of minimum reporting sets of patient characteristics in epidemiological research: a methodological systematic review"

Full search strategy of systematic review

- Phrases within 1 set are connected by OR
- Set A, set B, set C are connected by AND

| **Set A** | **Set B** | **Set C** | **NOT** | **Filter** |
| --- | --- | --- | --- | --- |
| baseline characteristic* [Title/Abstract] | standardized set* [Title/Abstract] | Consensus [Title/Abstract] | "clinical trial" [Publication Type] | From 2001 |
| patient characteristic* [Title/Abstract] | standardised set* [Title/Abstract] | delphi [Title/Abstract] | "randomized controlled trial" [Publication Type] | English |
| patients' characteristic* [Title/Abstract] | minimum set* [Title/Abstract] (no plural) | conceptual framework* [Title/Abstract] |  |  |
| patient-related characteristic* [Title/Abstract] | minimum reporting set* [Title/Abstract] (no plural) | conceptual analysis [Title/Abstract] |  |  |
| patient-related feature* [Title/Abstract] | standard*[Title/Abstract] | survey*[Title/Abstract] |  |  |
| patient feature* [Title/Abstract] | set[Title/Abstract] | interview[Title/Abstract] |  |  |
| patients’ feature* [Title/Abstract] | sets[Title/Abstract] | scoping review [Title/Abstract] |  |  |
| participant characteristic* [Title/Abstract] | reporting standard* [Title/Abstract] | systematic review [Title/Abstract] |  |  |
| participant-related characteristic* [Title/Abstract] | recommend* [Title/Abstract] | mixed method [Title/Abstract] |  |  |
| participants’ characteristic* [Title/Abstract] | suggest* [Title/Abstract] | literature review [Title/Abstract] |  |  |
| participant feature* [Title/Abstract] (no singular) |  | review [Title/Abstract] |  |  |
| prognostic factor* [Title/Abstract] |  | qualitative [Title/Abstract] |  |  |
| prognostic variable* [Title/Abstract] |  |  |  |  |
