## Appendix S2 for "Development of minimum reporting sets of patient characteristics in epidemiological research: a methodological systematic review"

Data extraction form of eligible studies

**Items**

Study ID

***General characteristics***

Publication year

Target medical conditions

Study purposes: Improving patient characteristics in

Did the study aimed to reach consensus?

Consensus-reaching method

Non-consensus method

Geographic scopes of study participants

***Methodological characteristics of Delphi studies***

1. *Participants*

Number of invited stakeholders by authors

Number of participated stakeholders

Response rate of each round well reported

Percentage of respondents of the last round

Type of stakeholders that participated in the study

Number of participants specified with each type of stakeholders

How were they chosen?

Authors reported how they ensured the representativeness of experts?

2. *Sources for identifying primary list of items*

3. *Delphi rounds*

3.1. Questionnaire rounds

Number of questionnaire rounds

Purposes of questionnaire rounds

Question formulation

Characteristics of rating scale?

Rating scale/ score well defined

Detailed questionnaire was available in the article or an appendix?

Material provided to the expert panel piloted in advance

Feedback that organizers sent each panel member

Were special techniques used to encourage participation? (For example: reminder or stamped addressed reply envelopes)

3.2. In-person/teleconference meeting rounds

Number of meeting rounds

Purposes of meeting round

Rating scale formulation

Rating scale/ score well defined

Criteria for selecting meeting participants

If modified Delphi procedure, when did the meeting occur?

3.3. Others

Additional items allowed to be added between rounds

4. *Consensus definition and attainment*

Criteria for selecting/dropping items at each round

Reason to terminate Delphi process

5. *Quality assessment criteria (proposed by Banno et al, 2019)*

1. Were criteria for participants reproducible? (Y/N) The method to select and exclude participants is stated. Number and type of participant subgroups (e.g., patients, generalists, and experts) are needed.

2. Was the number of rounds to be performed stated? (Y/N) We will categorize this as ‘Yes’ when the number of rounds is stated in the methods or results. We will categorize this as ‘Yes’ when researchers report the actual number of Delphi rounds in the results.

3. Were criteria for dropping items clear? (Y/N/NA) The prespecified criteria for dropping items at each round are reported.

4. Are stopping criteria, other than rounds, specified? (Y/N) The prespecified criteria for stopping the Delphi process, other than a statement of the number of rounds, are reported. For example, the prespecified criteria are related to the consensus or stability of responses

Total score

***Characteristics of the final lists of CPCS***

Flow chart or description of items flow

List of items reported

Number of items in the final list

Items in the final list defined

Measurement of non-obvious items in the final list

Were variables ranked?

Ranking of items in the final list

If ordinal variables were included, were levels of variables specified?

***Type of included variables/ characteristics***
