## Appendix S3 for "Development of minimum reporting sets of patient characteristics in epidemiological research: a methodological systematic review"

S3.1.Methodological characteristics of studies using Delphi consensus approaches (N=16)

| **Methodological characteristics** | **N** | **%** |
| --- | --- | --- |
| ***1. Participants*** |  |  |
| Number of invited individuals, median [IQR] | 62 [30.5–95] | |
| Number of individuals taking part in the study (i.e., participants), median [IQR] | 30 [24–60] | |
| Response rate of each round well reported | 12 | 75.0 |
| Response rate (%) of the last round, median [IQR] | 75.4 [56.0–100] | |
| Participant selection/exclusion based on ^a^: |  |  |
| - Renown, publishing, or expertise level | 9 | 56.3 |
| - Membership of a professional or patient organization | 6 | 37.5 |
| - Recommendations of other experts | 4 | 25.0 |
| - Years of experience | 2 | 12.5 |
| - Not reported | 3 | 18.8 |
| ***2. Methods to construct the primary list of items ^a^*** |  |  |
| - Systematic and/or scoping literature reviews | 8 | 50.0 |
| - Narrative and other non-systematic literature reviews | 6 | 37.5 |
| - Existing medical records or registry data forms | 5 | 31.3 |
| - Opinions of experts | 3 | 18.8 |
| - Unclear | 2 | 12.5 |
| ***3. Delphi rounds*** |  |  |
| ***3.1. Questionnaire rounds (in all 16 Delphi studies)*** |  |  |
| Purposes^a^ |  |  |
| - Rating items by a rating scale | 16 | 100.0 |
| - Collecting qualitative feedbacks^b^ | 10 | 62.5 |
| - Revising and approving final results | 1 | 6.3 |
| - Generating preliminary items | 1 | 6.3 |
| Material provided to the expert panel piloted in advance | 2 | 12.5 |
| Feedback that organizers sent to each panel member ^a^ |  |  |
| - Statistical summaries of previous round results | 8 | 50.0 |
| - Comments from panel members | 5 | 31.3 |
| - Not reported | 6 | 37.5 |
| Content of the questionnaires publicly accessible | 3 | 18.8 |
| Specific methods used to encourage participants to complete the questionnaires^c^ | 7 | 43.8 |
| ***3.2. In-person/teleconference meeting rounds (in 8 modified Delphi studies)*** | | |
| Purposes |  |  |
| - Discussing and rating items | 5 | 31.3 |
| - Discussing, rating and adding new item(s) | 1 | 6.3 |
| - Review the final results by ranking items | 1 | 6.3 |
| - Unclear | 1 | 6.3 |
| Participants invited to meetings |  |  |
| - All invited | 2 | 12.5 |
| - Selectively invited | 4 | 25.0 |
| - Not reported | 2 | 12.5 |
| Timing of meetings |  |  |
| - Before the first questionnaire round | 1 | 6.3 |
| - Between questionnaire rounds | 3 | 18.8 |
| - After the last questionnaire round | 4 | 25.0 |
| ***3.3 Others*** |  |  |
| Additional items allowed to be added between rounds | 11 | 68.8 |
| ***5. Quality score*** |  |  |
| - 1 point | 3 | 18.8 |
| - 2 points | 4 | 25.0 |
| - 3 points | 9 | 56.3 |
| ^a^ Each study may be classified in more than one category; ^b^ Including expert comments and/or suggestions for adding new items; ^c^By email reminders or voucher; IQR: interquartile range. | | |

S3.2. Quality score for each Delphi method conducted paper (proposed by *Banno et al*, 2019)

| **Study**  **(First author, published year, reference)** | **Quality assessment criteria ^a^** | | | | **Total score** |
| --- | --- | --- | --- | --- | --- |
|  | ***1. Were criteria for participants reproducible? (Yes or No)*** | ***2. Was the number of rounds to be performed stated? (Yes or No)*** | ***3. Were criteria for dropping items clear? (Yes, No or Not applicable)*** | ***4. Are stopping criteria, other than rounds, specified? (Yes or No)*** |  |
| Behrendt, 2018 | 0 | 1 | 0 | 0 | 1 |
| Cadihac, 2021 | 1 | 1 | 0 | 0 | 2 |
| Damhius, 2020 | 0 | 1 | 1 | 0 | 2 |
| Spronk, 2020 | 0 | 1 | 1 | 0 | 2 |
| Veer, 2018 | 1 | 1 | 1 | 0 | 3 |
| Wildi, 2013 | 1 | 1 | 1 | 0 | 3 |
| Lux, 2004 | 0 | 1 | 0 | 0 | 1 |
| Kendra Rio, 2019 | 1 | 1 | 1 | 0 | 3 |
| Goey, 2018 | 1 | 1 | 1 | 0 | 3 |
| Klimstra, 2010 | 1 | 1 | 1 | 0 | 3 |
| Ahmadi, 2015 | 0 | 1 | 0 | 0 | 1 |
| Khalil, 2019 | 1 | 1 | 1 | 0 | 3 |
| Lee, 2022 | 1 | 1 | 1 | 0 | 3 |
| Hamed, 2022 | 1 | 1 | 1 | 0 | 3 |
| Leonardi, 2022 | 0 | 1 | 1 | 0 | 2 |
| Prorok, 2022 | 1 | 1 | 0 | 1 | 3 |
| *Note: ^a^ Point given based on each criteria response: 1: Yes or NA (not applicable), 0: No.* | | | | | |

**S5.3.** Categories criteria for selecting/dropping items at each round of each study

| **Categories** | **Criteria for selecting/dropping items at each round** (Main author, publication year of quoted paper) |
| --- | --- |
| Pre-defined cut-off(s) of % of participants voting a certain rating level(s) | - Discussed items were added to the minimum data set if 80% of the experts supported the variable. (Behrendt 2018) - Items with an agreement level *(% of response at 4 or 5 points*) of less than 50% were identified for exclusion, between 50 and 79% required additional review, and 80% or more were accepted. (Cadihac 2021) - Baseline characteristics were presented to confirm inclusion when >70% of the experts scored a characteristic Likert score of 5. […] The consensus was defined with a pre-set level of agreement of [...] three out of three participants (100%) in the procedure to finalize the MRS. (Damhius 2020) - Baseline [...] characteristics that received more than 50% of votes from the expert panel were included in the mandatory set. (Veer 2018) - the predetermined threshold for consensus of >70% agreement or disagreement following the online process. (Kendra Rio 2019) - variables rated ‘very important’ by 67% of the experts were included in the recommended set. Variables rated ‘not important’ by 50% of the experts were excluded. [...] variables rated ‘very important’ by 67% of the experts were included in the recommended set. Remaining variables were incorporated in the suggested set, except variables rated ‘not important’ by 33% of the experts. Variables not fulfilling the criteria to be included in either set were excluded. (Goey 2018) - Consensus was defined to be achieved if at least 80% of the participants voted “yes” or “no”. (Klimstra 2010) - A predefined cut-off level of 70% agreement was used to define consensus for these questions. This meant that, [...], 70% of respondents should select ‘yes’. [...]. A cut-off level of 70% agreement was chosen to define consensus for these questions. (Khalil 2019) - Descriptors rated 7–9 by at least 70 per cent of clinicians were removed from subsequent rounds and formed a list of consensus descriptors to be discussed in the consensus meeting. Descriptors rated 1–3 by at least 70 per cent of clinicians were removed from the longlist of descriptors and not included in subsequent rounds. […] A threshold of 80 per cent agreement was required for any suggestion to be accepted. [...] The research team determined that an agreement threshold of 80 per cent was needed for inclusion. (Lee 2022) - The more stringent threshold was a rating of 4 or 5 by ≥80% of participants (threshold 2, preregistered), and the less-stringent threshold was a rating of ≥3 by ≥70% of participants (threshold 1). (Hamed 2022) - 1. Consensus that a data element/ outcome is important for a core domain set: ≥ 80% of participants in all groups scored the item as "critically important to include in a core set" (score 7 to 9) and < = 10% score as 1–3; these items are acknowledged in subsequent rounds as having met criteria for importance to a core set and held for final round discussion. 2. Consensus that a data element/ outcome will NOT be included: ≥ 50% of participants in all groups scored the item as of "limited importance" (score 1 to 6); these items are dropped from Delphi and are not to be part of core set. 3. Dissensus but important to one group: 80% + participants in one of our groups score items as critically important for a core set (score 7 to 9); data element/ outcome continues on to next round as having no consensus yet; if data element/ outcome does not reach consensus level at end of Delphi, but still important to one group, it will be held for final round discussion, 4. No consensus: All other results; data element/ outcome continues to next round as having no consensus yet. If data element/ outcome does not achieve consensus by last round, and no groups have supported it ≥ 80%, then data element/ outcome is not endorsed for core set (Prorok 2022) - In some cases, the experts were simply asked to agree or disagree with a proposed change. For this type of question, a minimum of 70% of experts needed to agree to be considered consensus (Leonardi 2022) |
| Pre-defined cut-off(s) of a median score on a rating scale | - The predefined cut-off for inclusion was a median Likert score of 5 (Damhius 2020) - we only included characteristics that had been rated a median of seven or above by the expert panel. (Wildi 2013) - The predefined cut-off for inclusion of variables in the MRS was a median score of 5 on the Likert scale. (Khalil 2019) |
| Composite criteria | - Inclusion criteria in the global common data set: (1) median score of 5 or 6; (2) more than 70 percent of the panel scoring a 5 or 6. (Spronk 2020) - The features that were planned for inclusion into the final round of the Delphi process required a median Likert score of 5 (equivalent to “agree”) and a proportion of experts above 90% that selected a Likert score of 5 to 7. Features that were planned for re-evaluation in round 2 required a median Likert score of 5 and a proportion of experts between 60% and 90% that selected a Likert score of 5 to 7 and not more than 15% of experts selecting a Likert score of 1 to 3. Features that had a median Likert score of <5, or had >15% of experts selecting a Likert score of 1 to 3 were not considered for the next round. (Leonardi 2022) |
| Unclear/ unclassified criteria | - Items repeatedly (*not defined*) rated with “strongly agree” or “agree” were recommended for the minimum data set. Items repeatedly rated with “strongly disagree” or “disagree” were eliminated from consideration (Behrendt 2018) - data elements with agreement levels (*not defined*) less than 50% were excluded at the first round, 50 to 75% agreement levels entered the second round, and agreement levels more than 75% were accepted. [...] In the second round, an agreement level of 75% was considered on each data element (Ahmadi 2015) - The data element or outcome in each domain which received the most rankings as the top data element or outcome formed part of the primary core set, while the data element or outcome in each domain receiving the most rankings as the second most important data element or outcomes formed part of the secondary core set. (Prorok 2022) |
