## Appendix S5 for "Development of minimum reporting sets of patient characteristics in epidemiological research: a methodological systematic review": Appendix S5. Cluster plot code.Rmd

````
---
title: "CBCS-plot"
author: "Trang Tu"
date: "2/20/2022"
output: html_document
---

```{r setup, include=FALSE}
knitr::opts_chunk$set(echo = TRUE)
```

```{r}
library(curl)
DB1 <- read.csv(curl("https://raw.githubusercontent.com/trangtph/cbcs/main/cluster_data.csv") )
```


```{r, results='hide',message=FALSE, warning=FALSE, echo=FALSE}
setwd("/Users/tranghien/Desktop/Mediation Review/CBCS Systematic Review/Result Report/Tree-Figure")
```

```{r}
library(tidyverse)
library(data.table)
```

```{r}

domains1 = c("Demographic factors", "Clinical factors", "Patient history factors", "Socioeconomic factors", "Healthcare setting factors")

#Names of the tools (n=21)
tools1 = c("Behrendt 2018", "Cadihac 2021", "Damhius 2020", "Driessen 2016","Spronk 2020","Veer 2018","Wildi 2013","Lux 2004","Osooli 2015","Kendra Rio 2019","Goey 2018","Jason 2012","Klimstra 2010","Kwakkel 2017","Storrow 2012","Jones 2018","Khalil 2019","Lee 2022", "Hamed 2022", "Leonardi 2022", "Prorok 2022")
```

```{r}
Q1 <- DB1 %>% select(Demographic.factors., Clinical.factors., Patient.history.factors., Socioeconomic.factors., Healthcare.setting.factors.)
Q1$Demographic.factors. <- as.numeric(Q1$Demographic.factors.)
Q1$Clinical.factors. <- as.numeric(Q1$Clinical.factors.)
Q1$Patient.history.factors. <- as.numeric(Q1$Patient.history.factors.)
Q1$Socioeconomic.factors. <- as.numeric(Q1$Socioeconomic.factors.)
Q1$Healthcare.setting.factors. <- as.numeric(Q1$Healthcare.setting.factors.)
setDT(Q1)
filas1=apply(Q1,1,sum)
filas1
T11 <- diag(1/filas1) %*% as.matrix(Q1)
T11
d11 = dist(T11) #Euclidean distance between two vectors, A and=√Σ(Ai-Bi)2
H1 = hclust(d11) #Hierachical clustering: group a set of data points into subsets or clusters: to create clusters that are coherent internally, but clearly different from each other externally. 
#Complete-linkage: calculates the maximum distance between clusters before merging.
H1$order
plot(H1)
```

```{r}
T1 = as.data.frame(t(Q1)) #number of items in each category for 21 studies
colnames(T1) <- c(1:21)
T1
PAL=c(rgb(251,154,153, max=255), rgb(178,223,138, max=255), rgb(31,120,180, max=255), rgb(227,26,28, max=255),
      rgb(51,160,44, max=255)) # creat RGB pallets? 
```


```{r}
sector = function(Cent=c(0,0), A, R, bor=NA, color, dens=NULL, ang=90) {
  if (abs(R[1]-R[2]) < 1e-8) return() #absolute values -> returns the last output of the function
  if (abs(A[1]-A[2]) < 1e-8) return()
  X = c(R[1]*cos(A[1]), R[2]*cos(A[1]))
  Y = c(R[1]*sin(A[1]), R[2]*sin(A[1]))
  a = seq(A[1], A[2], len=40) #create sequence from A1 to A2 with length = 40
  X = c(X, R[2]*cos(a), R[1]*cos(A[2]))
  Y = c(Y, R[2]*sin(a), R[1]*sin(A[2]))
  a = seq(A[2], A[1], len=40) 
  X = c(X, R[1]*cos(a))
  Y = c(Y, R[1]*sin(a))
  X = X + Cent[1]
  Y = Y + Cent[2]
  polygon(X, Y, border=bor, col=color, density=dens, angle=ang) #plot a polygon between specified points 
}
```

Clusters of tool
```{r}
L = list(c(13,20,7,16), c(21,8,5,6), list(c(1, 17), c(4, 15, 9, 19)), list(c(2), c(14,3,12,18,10,11)))
```

```{r}
draw_tool = function(r, a2) {
  t = T1[,r] #the rth column of the table T above
  p = c(0, cumsum(rev(t))/sum(t)) #rev(): reverse order of the columns/ vector
  s = sqrt(R1^2 + 2*(1-p)/x/K) 
  for (m in 1:an) {
    mm = an-m+1
    sector(A=c(a2-x/2, a2+x/2), R=s[m:(m+1)], color=pal[mm])
  }
  p = P*s[1]
  b = ifelse(a2<0 & a2>-pi, a2+pi, a2)
  text(p*cos(a2), p*sin(a2), srt=180*(b-pi/2)/pi, lab=tools1[r],cex=1.7) 
}
```

```{r}
#Save the figure as png
png('cluster3.png', width=1500, height=1500)
par(mar=c(1,1,0,0)) #define margin around the plot
v = 1.7
plot(NA, xlim=c(-v,v), ylim=c(-v,v), xlab='', ylab='', axes=FALSE)
an = dim(T1)[1] #number of rows in table T
bn = dim(T1)[2] #number of columns in table T

pal = PAL
P = 1.05
R1 = 0.45
R2 = 1.5
Ang.0 = 1.570795
Ang.1 = -pi-pi/10
gap = 0.12
x = (Ang.1 - Ang.0)/((bn-1 + 5)*gap + bn)  
K = 2/x/(R2^2-R1^2)
lev = c(0.25, 0.5, 0.75)
for (i in 1:length(lev)) {
  s = sqrt(R1^2 + 2*(1-lev[i])/x/K)
  a = Ang.0-(x/4)
  sector(A=c(Ang.0-x/5, Ang.1+2*x/5), R=c(0.99*s, s), color='grey')
  text(s*cos(a), s*sin(a), lev[i], col='grey2', pos=2, cex=1.25)
}

#Legend of the figure 
legend(-v, 0.94*v, legend=domains1, fill=pal, density=NA, bty='n', cex=2.25) #bty: the type of box to be drawn around the legend.
w = Ang.0
clu = rep(NA, 6); kl = 0
for (i in 1:length(L)) {
  LL = L[[i]]
  if (is.list(LL)) {
    # line from center to center of tools in list (i,*), at level 1
    n = length(unlist(LL))
    a = w + n/2*x + (n-1)/2*x*gap
    segments(0, 0, R1/3*cos(a), R1/3*sin(a), lwd=4, col="royalblue3")
    w1 = 0
    for (j in 1:length(LL)) {
      # line from center of tools in list (i,*) to center of tools in list (i,j), at level 2
      l = unlist(LL[[j]])
      n1 = length(l)
      a1 = w + w1 + n1/2*x + (n1-1)/2*x*gap
      kl=kl+1; clu[kl]=a1
      segments(R1/3*cos(a), R1/3*sin(a), 2*R1/3*cos(a1), 2*R1/3*sin(a1), lwd=4, col="royalblue3")
      a2 = w + w1 + x/2    
      for (k in 1:n1) {
        # line from ... (i,j) to each tool in this list at level 3 (R1)
        segments(2*R1/3*cos(a1), 2*R1/3*sin(a1), R1*cos(a2), R1*sin(a2), lwd=4, col="royalblue3")
        draw_tool(l[k], a2)  
        a2 = a2 + x*(1+gap)
      }
      sector(A=c(w+w1, a2-x*(0.5+gap)), R=c(1.115,1.165)*R2, color='papayawhip')
      w1 = w1 + n1*x*(1+gap) + x*gap
    }
  } else {  
    n = length(LL)
    a = w + n/2*x + (n-1)/2*x*gap
    kl=kl+1; clu[kl]=a
    segments(0, 0, 2*R1/3*cos(a), 2*R1/3*sin(a), lwd=4, col="royalblue3")
    n1 = n
    a2 = w + x/2    
    for (k in 1:n1) {
      # line  to each tool in this list at level 3 (R1)
      segments(2*R1/3*cos(a), 2*R1/3*sin(a), R1*cos(a2), R1*sin(a2), lwd=4, col="royalblue3")
      draw_tool(LL[k], a2)  
      a2 = a2 + x*(1+gap)
    }
    sector(A=c(w, a2-x*(0.5+gap)), R=c(1.115,1.165)*R2, color='papayawhip')
    w1 = a2-w-x/2+x*gap
  }
  w = w + w1 + 1.5*x*gap
}
for (i in 1:6) {
  a = clu[i]
  b = ifelse(a<0 & a>-pi, a+pi, a)
  tx = sprintf("#%d", i)
  text(1.14*R2*cos(a), 1.14*R2*sin(a), srt=180*(b-pi/2)/pi, lab=tx, cex=2.5)
}
dev.off() #closes the specified plot
```

```{r}

```

```{r}

```

```{r}

```

```{r}

```

```{r}

```
````
